# Uncovering the linguistic characteristics of psychotherapy: a computational approach to measure therapist language timing, responsiveness, and consistency

**DOI:** 10.1101/2022.04.24.22274227

**Authors:** Adam S Miner, Scott L Fleming, Albert Haque, Jason A Fries, Tim Althoff, Denise E Wilfley, W. Stewart Agras, Arnold Milstein, Jeff Hancock, Steven M Ash, Shannon Wiltsey Stirman, Bruce A. Arnow, Nigam H. Shah

**Affiliations:** Department of Psychiatry and Behavioral Sciences, Stanford University, Stanford, CA, USA; Center for Biomedical Informatics Research, Stanford University, Stanford, CA, USA; Department of Biomedical Data Science, Stanford University, Stanford, CA, USA; Department of Computer Science, Stanford University, Stanford, CA, USA; Allen School of Computer Science & Engineering, University of Washington, Seattle, WA, USA; Departments of Psychiatry, Medicine, Pediatrics, and Psychological & Brain Sciences, Washington University in St. Louis, St. Louis, MO, USA; Clinical Excellence Research Center, Stanford University, Stanford, CA, USA; Department of Communication, Stanford University, Stanford, USA; VA Palo Alto Health Care System; Division of Primary Care and Population Health, Stanford University School of Medicine; National Center for Posttraumatic Stress Disorders, Dissemination and Training Division, VA Palo Alto Healthcare System, Menlo Park, California, United States; Technology and Digital Solutions, Stanford Healthcare

**Author notes:** Co-first authors (Adam S Miner, Scott L Fleming).

## Abstract

Although individual psychotherapy is generally effective for a range of mental health conditions, little is known about the moment-to-moment language use of effective therapists. Increased access to computational power, coupled with a rise in computer-mediated communication (telehealth), makes feasible the large-scale analyses of language use during psychotherapy. Transparent methodological approaches are lacking, however. Here we present novel methods to increase the efficiency of efforts to examine language use in psychotherapy. We evaluate three important aspects of therapist language use - timing, responsiveness, and consistency - across five clinically relevant language domains: pronouns, time orientation, emotional polarity, therapist tactics, and paralinguistic style. We find therapist language is dynamic within sessions, responds to patient language, and relates to patient symptom diagnosis but not symptom severity. Our results demonstrate that analyzing therapist language at scale is feasible and may help answer longstanding questions about specific behaviors of effective therapists.

## Introduction

Individual psychotherapy is an effective treatment for a wide range of mental health conditions^1,2^.Two problems that have emerged in research on outcomes from psychotherapy are that: 1) for some disorders, there is little evidence that one specific form of psychotherapy is superior to another even when hypothesized change mechanisms differ^3^, and 2) some therapists consistently achieve better outcomes than others (i.e., therapist effects), but it is unclear what individual therapists may be doing that accounts for these effects^4–7^. Studies of the psychotherapy process attempt to understand what happens during the sessions that may explain patient improvement^8^. The chief method used since the 1950s to evaluate therapist behavior in therapy sessions is to have trained humans identify clinically meaningful therapist utterances in transcripts, and draw conclusions based on observed patterns^9–12^. Although useful, relying solely on human inspection of transcripts is not likely to meet demands for improved reproducibility and scalability in psychotherapy process research^11,13–17^.

Computational approaches using natural language processing offer the potential to move past human limits of attention and reproducibility^11,18–22^. Improvements in computational power, the growing ease of recording and transcribing therapy sessions, and a shift to computer-mediated communication in healthcare (i.e., telehealth) make this feasible^11,14,23,24^. Early work is promising, but does not yet translate to best practices for improved patient outcomes or provide a clear direction for therapist training^11,19,25–28^. Methodological improvements are needed to bridge divisions between theoretical schools of thought (e.g., Cognitive Behavioral, Interpersonal, Psychodynamic, Counseling) as to which therapist language patterns correlate with favorable therapy outcomes^4,6,13,15,20,24,29,30^. If known, the linguistic behavior of successful therapists may inform targeted clinical trials to test causality and implementability, subsequently improving clinician training.

A fundamental tenet of psychotherapy is that therapists expose patients to language that may be helpful (e.g., emotional validation) and avoid language that may be harmful (e.g., shaming). Therapist language should be well timed and appropriate for the specific moment. Nevertheless, the specific timing, frequency, and reactivity of therapist utterances is difficult to scrutinize systematically without human inspection^13^. Difficulties are multifaceted, with key limitations being theoretical (i.e., disagreement about mechanisms of change), technical (i.e., lack of validated tools for language measurement), and practical (i.e., lack of clinically meaningful datasets). This work primarily addresses the technical limitations of language analysis in psychotherapy. Here we present a three-phase approach that measures therapist language by building on prior theoretical, methodological, and clinical insights: **Phase 1** - To identify *a priori* language features of interest, we generate a non-exhaustive list of clinically relevant language features. **Phase 2** - To observe the natural occurrence of language features identified in Phase 1, we describe the underlying structure of therapy focusing on <u>timing</u>, <u>responsiveness</u>, and <u>consistency</u>. **Phase 3** - To demonstrate the potential for clinical utility, we evaluate the relationship between therapist language and patient symptom severity and diagnosis.

Many forms of therapy exist, along with an abundance of theoretically and practically motivated therapist approaches. Thus, we suggest a reasonable but non-exhaustive list of domain-focused concepts that balance face-validity and technical implementability using modern linguistic and statistical approaches. We posit, based on prior research and clinical judgment, that five clusters of language features may be clinically important across theoretical orientations, meriting close inspection. We limit our focus to characterizations of human language most amenable to machine learning, and that may correlate with favorable patient improvement. We acknowledge that other modern sensing technologies will allow for more rich characterization of human interaction such as facial expressions, body movement, and voice tone that may also be related to therapy outcomes^31^.

The five feature clusters we seek to describe are: pronouns, time orientation, emotional polarity, therapist tactics, and paralinguistic style. **Pronouns** (e.g., I, me, you, them) reflect internal psychological attention^25,32,33^. Measuring the relative frequency of self-focused pronouns (i.e., I, me, my) and other-focused pronouns (e.g., you, your, they) has demonstrated theoretical and practical value in psychological research^32,34,35^. **Time orientation** is a longstanding focus of psychotherapy. Some theoretical orientations advise therapists to focus on past experiences (e.g., early childhood), while some encourage focus on the present^36–39^. **Emotions** are important in most clinical psychology theoretical orientations^36,37,40–42^. There is strong disagreement, however, on how to represent and measure polarity and emotionality in clinical contexts^43–45^. **Therapist tactics** are used to help develop a therapeutic relationship and engender patient change, including statements that demonstrate understanding^4,11^. **Paralinguistics** refers to the way words are said, not the words themselves, for example, rate of speech^46,47^. Based on prior work, these language-focused constructs are theoretically important, but poorly measured in psychotherapy. Although a full review of the theoretical importance and practical application of these clusters is beyond the scope of this work, we briefly summarize each feature in our Methods (Phase 1: Feature generation).

Uncovering modifiable, therapist-focused interventions that are associated with patient improvement is a key objective of therapy process research^4,8,13,15^. Our approach presents a systematic way to generate or evaluate hypotheses about psychotherapy process at scale. This study identifies potentially modifiable features of interest in psychotherapy (Phase 1), measures feature timing, responsiveness, and consistency (Phase 2), tests clinical usefulness (Phase 3), and shares methods to encourage critical peer review and collaboration.

## Results

Overall, our results surface linguistic nuance in psychotherapy that previously has not been directly measured. Therapist language <u>timing</u> is dynamic (Figure 1) and does not mirror the frequency of patient language consistently (Figure 2). Therapist language appears to be <u>responsive</u> to patient language for a number of clinically relevant language features (Figures 3, 4). For example, Figures 3 and 4 show that therapists decreased their rate of speech, as measured by words per second, in response to increases in the patient’s rate of speech, or vice versa (i.e., therapists significantly slowed their speech as patients increased theirs). Therapist language appears <u>consistent</u> across sessions: on average, within-therapist language patterns were significantly more similar than between-therapist language patterns. In relation to patient-focused characteristics, therapist language appears to be related to patient diagnosis: logistic regression models trained to classify diagnosis based on therapist language patterns performed significantly better than chance.

**Figure 1.**
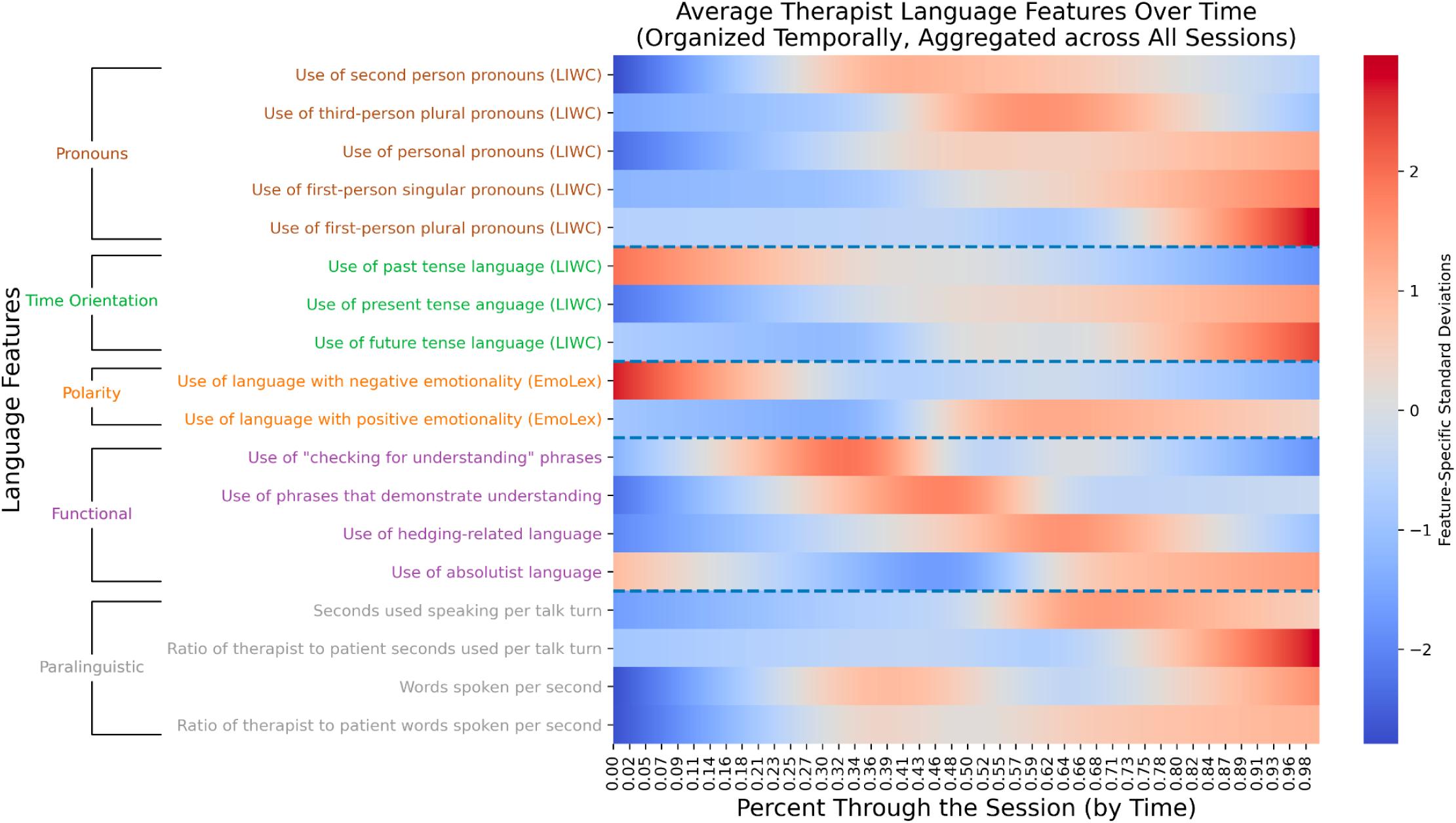
Therapist speech phase-dependence. The dynamic nature of therapist speech, grouped by language feature category. Figure 1 represents trends in therapist language over time after aggregating across therapists. LIWC = <u>L</u>inguistic <u>I</u>nquiry and <u>W</u>ord <u>C</u>ount, a dictionary-based lexicon that maps words and word stems to psychologically relevant categories. EmoLex = Word-<u>Emo</u>tion Association <u>Lex</u>icon, a list of English words mapped to crowdsourced sentiment annotations. We performed smoothing/interpolation between discrete points at the level of temporal quintiles using a natural cubic spline. See Figure 2 for per-feature examples of these trends viewed without smoothing.

**Figure 2.**
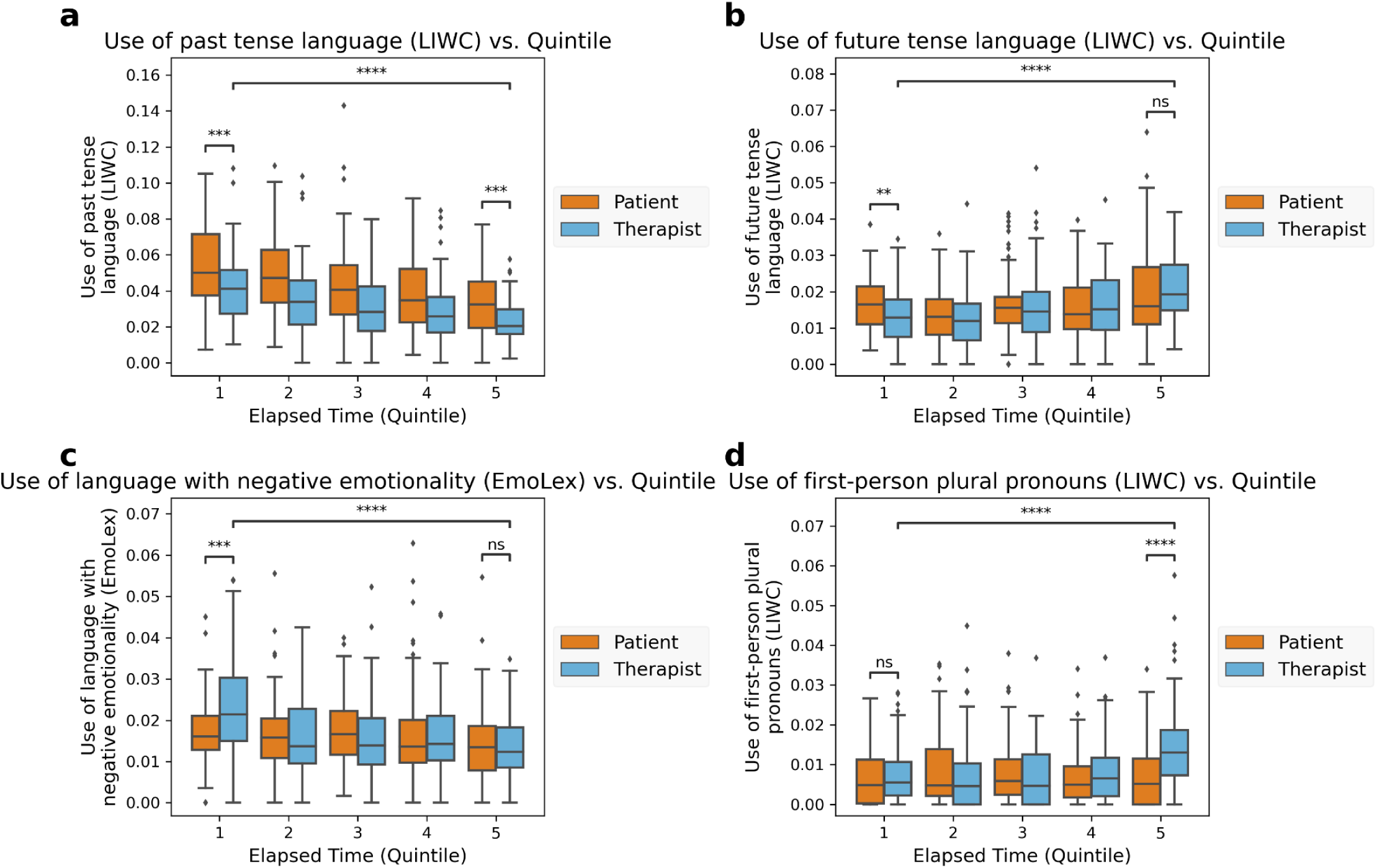
Therapist and patient language within-session changes. Quantitative assessment of changes in therapist language features over time, as well as within-quintile differences between patient and therapist language. All differences annotated asterisks (*) are significant at level α=0.05 after controlling for multiple hypothesis tests via the Benjamini-Hochberg procedure. p-value annotation: Non-significant (ns): 0.01 < p ≤ 1.0; *: 0.01 < p ≤ 0.05; **: 0.001 < p ≤ 0.01; ***: 0.0001 < p ≤ 0.001; ****: p ≤ 0.0001

**Figure 3.**
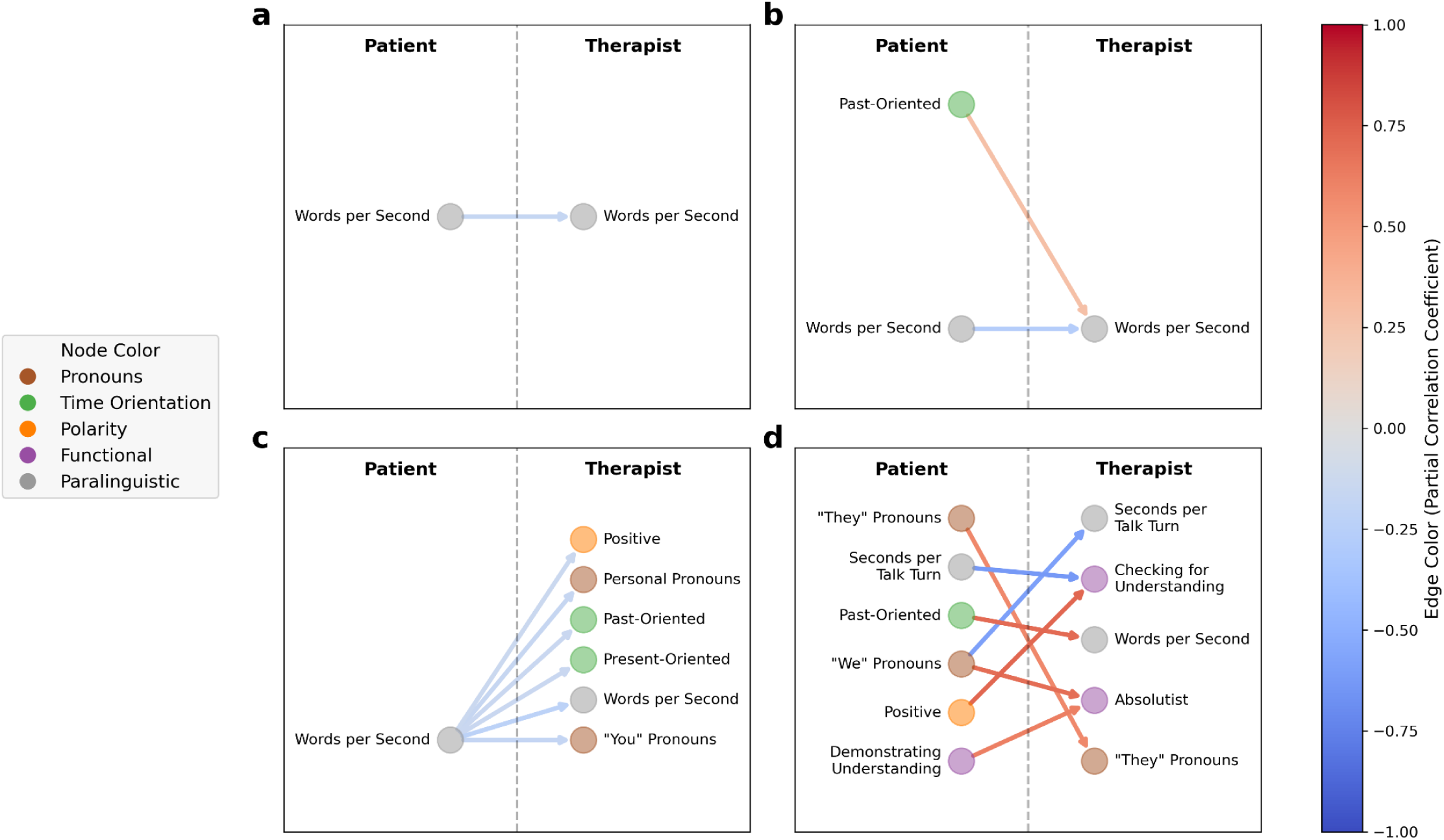
Therapist responsiveness patterns at the level of individual sessions. Illustration of significant directional associations between patient language and therapist language in four sessions, each representing a unique patient-therapist dyad. Language features are colored by feature group (see Table 2). Edges are colored according to the average partial correlation coefficient. Figure 3a illustrates an example of one patient-therapist dyad in which there was just one significant association: increases in patient rate of speech, as measured in words per second, were associated with decreases in therapist rate of speech, and vice versa. Figure 3d highlights a patient-therapist dyad with varied significant associations: increased patient use of third-person plural pronouns (‘“They” Pronouns’) drove increased therapist use of third-person plural pronouns (‘“They” Pronouns’), increased use of positive language by the patient (“Positive”) was associated with increased use of checking for understanding phrases by the therapist (“Checking for Understanding”), etc. These are four of the 73 network diagrams produced, one for each session/patient-therapist dyad.

**Figure 4.**
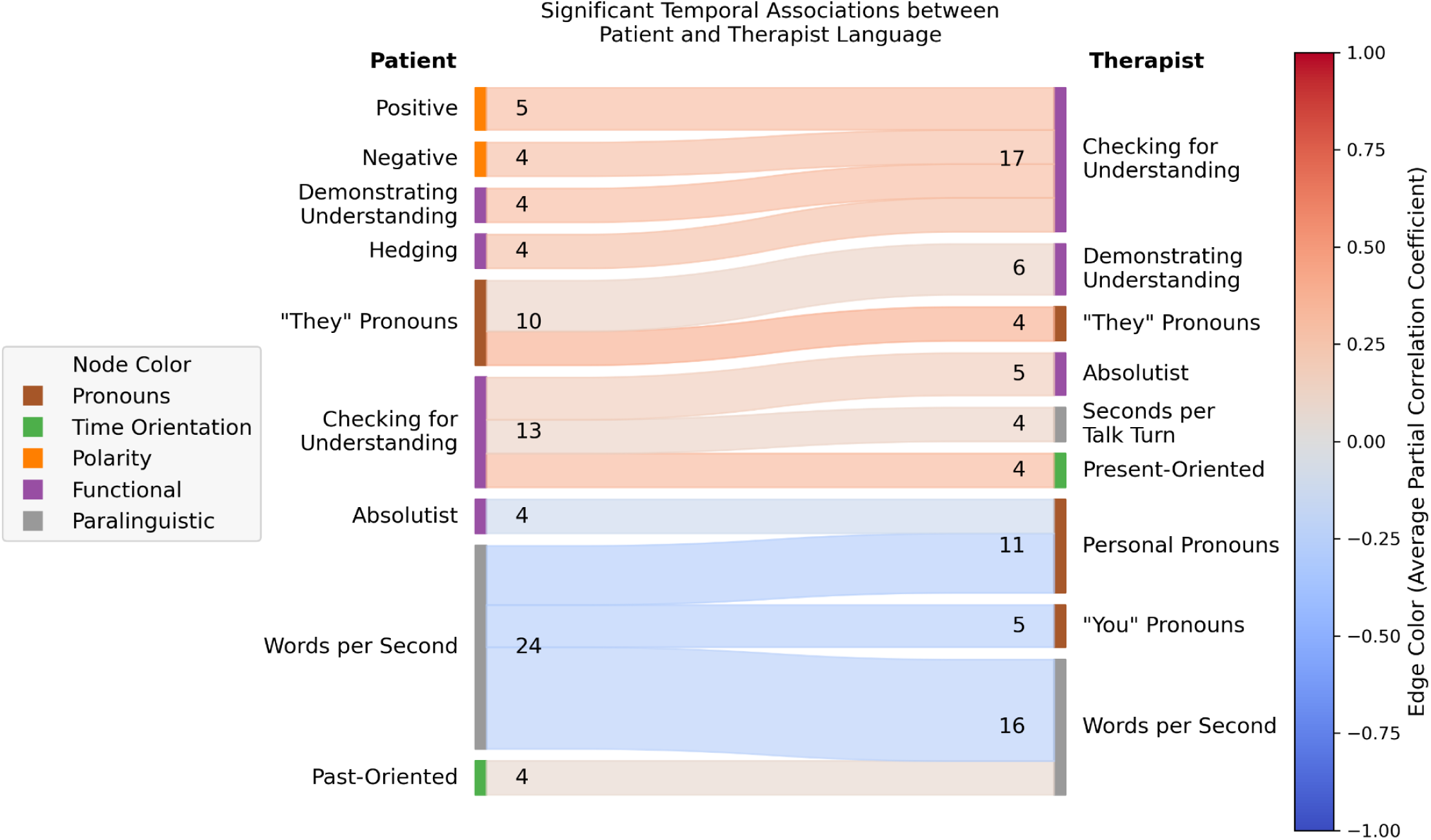
Therapist responsiveness patterns aggregated over all sessions. The number of times a particular type of association between patient language features and subsequent/accommodating therapist language features was found, across all sessions. Patient language features are on the left, therapist language features on the right. For the purposes of illustration, only associations that were found in at least 4 patient-therapist dyads are displayed (see Supplementary Figure 2 for a similar plot containing all significant associations). There were 72 such associations from 43 unique patient-therapist dyads, of which 24 involved changes in the patient’s rate of speech (“Words per Second”). Language features are colored by feature group (see Table 2). Edges are colored according to the average partial correlation coefficient amongst all patient-therapist dyads in which that association was found. For example, 12 patient-therapist dyads exhibited a significant negative association between patient rate of speech and therapist rate of speech, such that increases in the patient’s words per second (“Words per Second”) were associated with subsequent decreases in the therapist’s words per second (“Words per Second”) and/or vice versa (i.e., decreases in the patient’s words per second were associated with subsequent increases in the therapist’s words per second).

### Study population

Therapy transcripts were created per protocol as part of a secondary analysis of a previously completed randomized controlled trial, conducted in the United States across 24 college counseling clinics from April 2013 and December 2016^14,48^. See [Miner et al., 2020]^14^ for details on transcription and sample selection. The demographic information of a subset of therapist-patient dyads, and their clinical information (diagnosis and symptom severity) is presented in Table 1. Patients were predominantly female (87%) and in their early 20s (median age, 21 years). Therapists were predominantly female (78%), and in their early 40s (median age, 41 years). Patient depressive symptom severity was mostly minimal to mild.

**Table 1.**
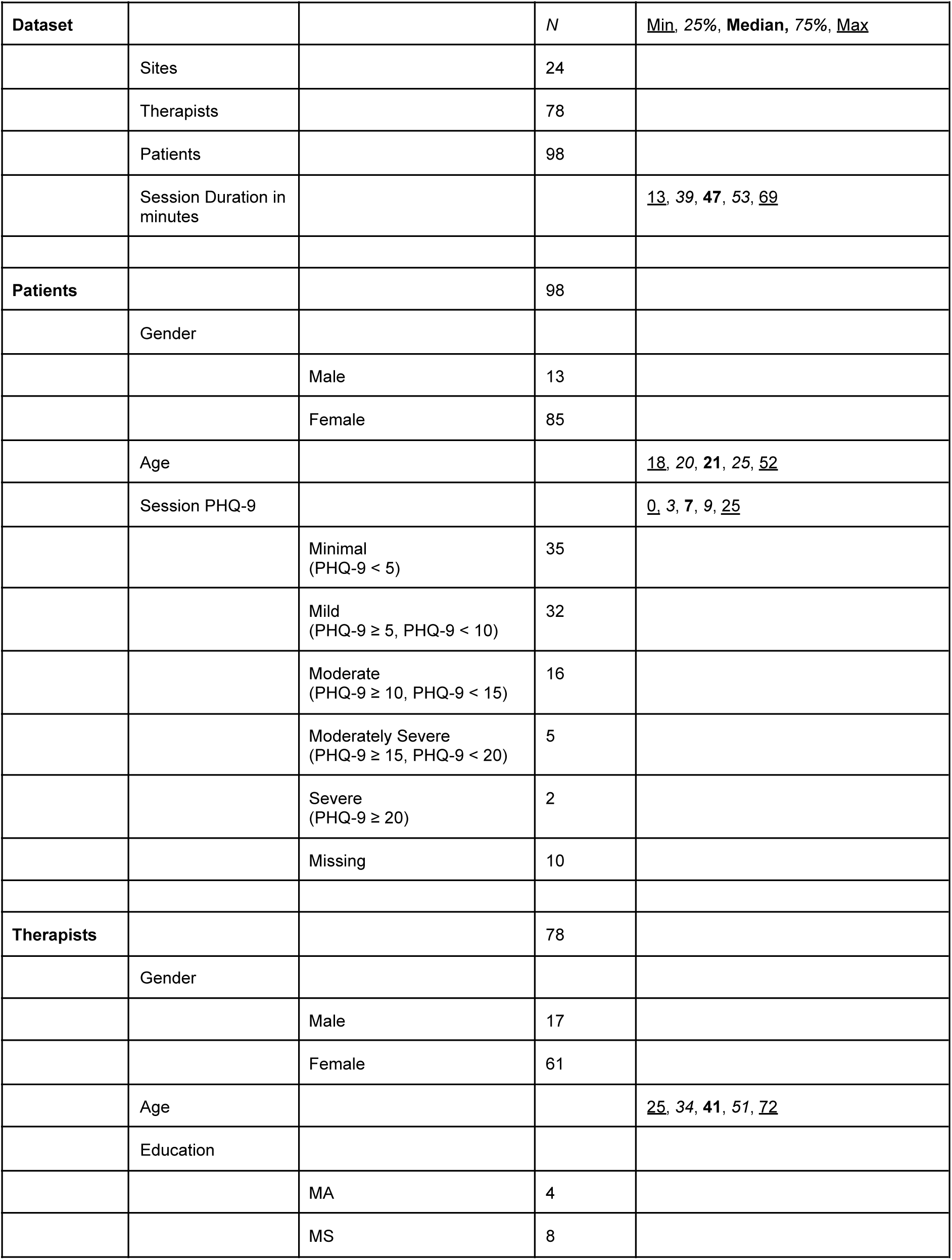
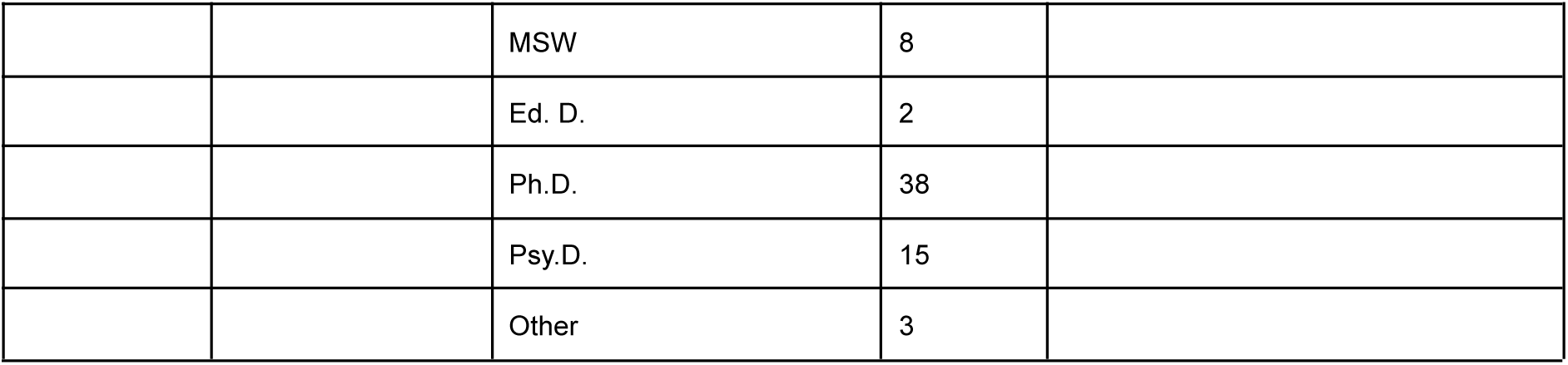
Clinician and patient demographic information

| Dataset |  |  | N | Min, 25%, Median, 75%, Max |
| --- | --- | --- | --- | --- |
|  | Sites |  | 24 |  |
|  | Therapists |  | 78 |  |
|  | Patients |  | 98 |  |
|  | Session Duration in minutes |  |  | <u>13</u>, 39, **47**, 53, <u>69</u> |
| **Patients** |  |  | 98 |  |
|  | Gender |  |  |  |
|  |  | Male | 13 |  |
|  |  | Female | 85 |  |
|  | Age |  |  | <u>18</u>, 20, **21**, 25, <u>52</u> |
|  | Session PHQ-9 |  |  | <u>0</u>, 3, 7, 9, <u>25</u> |
|  |  | Minimal (PHQ-9 < 5) | 35 |  |
|  |  | Mild (PHQ-9 ≥ 5, PHQ-9 < 10) | 32 |  |
|  |  | Moderate (PHQ-9 ≥ 10, PHQ-9 < 15) | 16 |  |
|  |  | Moderately Severe (PHQ-9 ≥ 15, PHQ-9 < 20) | 5 |  |
|  |  | Severe (PHQ-9 ≥ 20) | 2 |  |
|  |  | Missing | 10 |  |
| **Therapists** |  |  | 78 |  |
|  | Gender |  |  |  |
|  |  | Male | 17 |  |
|  |  | Female | 61 |  |
|  | Age |  |  | <u>25</u>, 34, **41**, 51, <u>72</u> |
|  | Education |  |  |  |
|  |  | MA | 4 |  |
|  |  | MS | 8 |  |
|  |  | MSW | 8 |  |
|  |  | Ed. D. | 2 |  |
|  |  | Ph.D. | 38 |  |
|  |  | Psy.D. | 15 |  |
|  |  | Other | 3 |  |

**Table 2.**
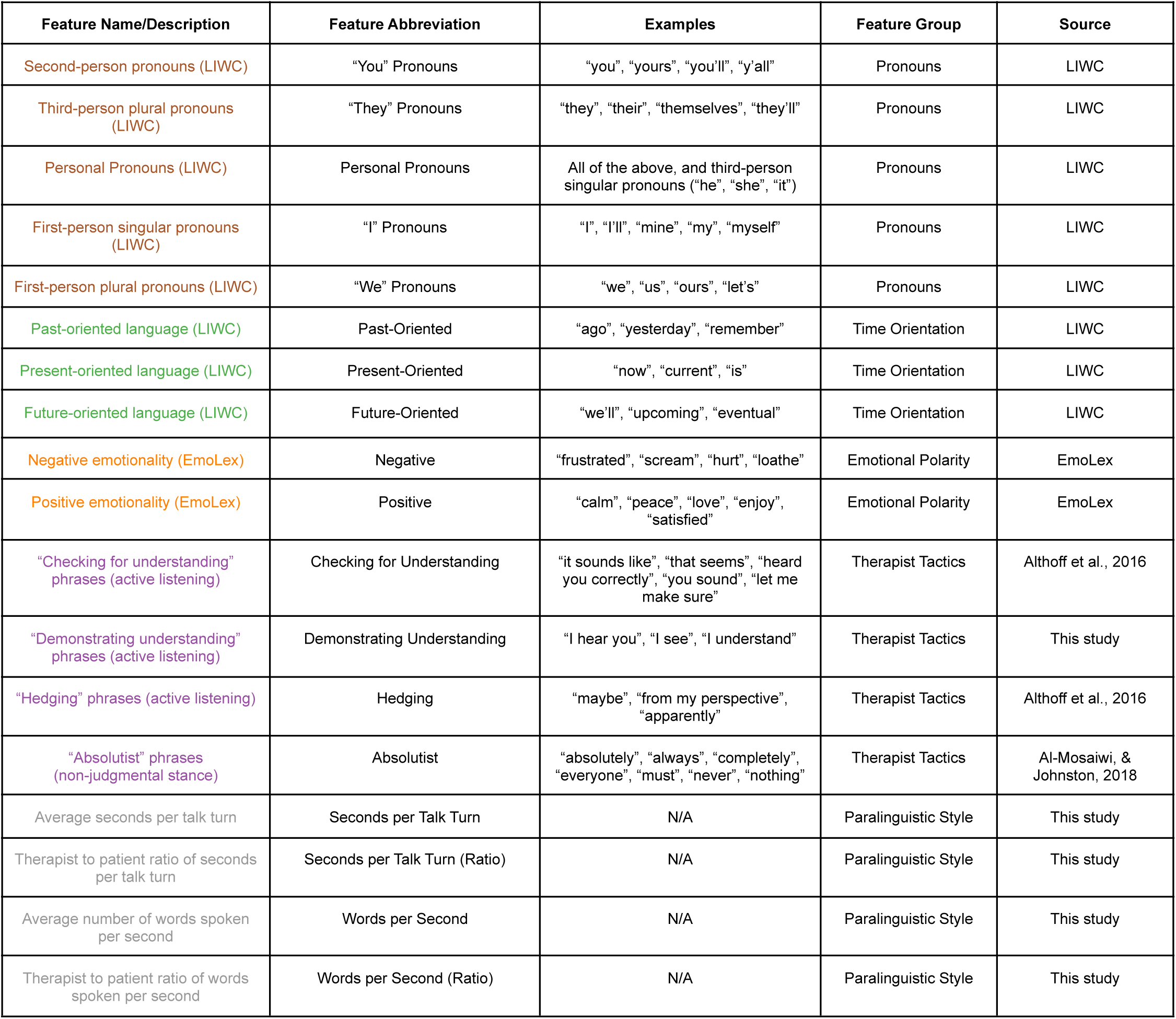
Summary of language features

| Feature Name/Description | Feature Abbreviation | Examples | Feature Group | Source |
| --- | --- | --- | --- | --- |
| Second-person pronouns (LIWC) | "You" Pronouns | "you", "yours", "you'll", "y'all" | Pronouns | LIWC |
| Third-person plural pronouns (LIWC) | "They" Pronouns | "they", "their", "themselves", "they'll" | Pronouns | LIWC |
| Personal Pronouns (LIWC) | Personal Pronouns | All of the above, and third-person singular pronouns ("he", "she", "it") | Pronouns | LIWC |
| First-person singular pronouns (LIWC) | "I" Pronouns | "I", "I'll", "mine", "my", "myself" | Pronouns | LIWC |
| First-person plural pronouns (LIWC) | "We" Pronouns | "we", "us", "ours", "let's" | Pronouns | LIWC |
| Past-oriented language (LIWC) | Past-Oriented | "ago", "yesterday", "remember" | Time Orientation | LIWC |
| Present-oriented language (LIWC) | Present-Oriented | "now", "current", "is" | Time Orientation | LIWC |
| Future-oriented language (LIWC) | Future-Oriented | "we'll", "upcoming", "eventual" | Time Orientation | LIWC |
| Negative emotionality (EmoLex) | Negative | "frustrated", "scream", "hurt", "loathe" | Emotional Polarity | EmoLex |
| Positive emotionality (EmoLex) | Positive | "calm", "peace", "love", "enjoy", "satisfied" | Emotional Polarity | EmoLex |
| "Checking for understanding" phrases (active listening) | Checking for Understanding | "it sounds like", "that seems", "heard you correctly", "you sound", "let me make sure" | Therapist Tactics | Althoff et al., 2016 |
| "Demonstrating understanding" phrases (active listening) | Demonstrating Understanding | "I hear you", "I see", "I understand" | Therapist Tactics | This study |
| "Hedging" phrases (active listening) | Hedging | "maybe", "from my perspective", "apparently" | Therapist Tactics | Althoff et al., 2016 |
| "Absolutist" phrases (non-judgmental stance) | Absolutist | "absolutely", "always", "completely", "everyone", "must", "never", "nothing" | Therapist Tactics | Al-Mosaiwi, & Johnston, 2018 |
| Average seconds per talk turn | Seconds per Talk Turn | N/A | Paralinguistic Style | This study |
| Therapist to patient ratio of seconds per talk turn | Seconds per Talk Turn (Ratio) | N/A | Paralinguistic Style | This study |
| Average number of words spoken per second | Words per Second | N/A | Paralinguistic Style | This study |
| Therapist to patient ratio of words spoken per second | Words per Second (Ratio) | N/A | Paralinguistic Style | This study |

### Therapist timing is dynamic

Here we evaluate therapist language timing. Therapists appear to use distinct types of language at specific points in the session (early vs. late feature frequency). Figure 1 presents normalized frequency over time of therapist language features. Supplementary Figure 1 shows individual therapists as examples. Figure 2 presents differences between therapist and patient language features over time for a subset of features.

Therapist speech changed significantly between the start and the end of the session. As illustrated in Figure 1 and Supplementary Table 1, relative to the first quintile of the session, therapists in the last quintile of the session used a smaller proportion of words with **negative emotionality** (0.0136 vs. 0.0227, *p*=3.97 × 10^−7^); a greater proportion of **present-focused words** (0.1697 vs. 0.1271, *p*=1.30 × 10^−15^) and **future-focused words** (0.2084 vs. 0.01314, *p*=2.46 × 10^−7^), but a smaller proportion of **past-focused words** (0.0231 vs. 0.0416, *p*=6.87^-11^); and a greater proportion of **personal pronouns** (0.1500 vs. 0.1182, *p*=3.86 × 10^−10^), including first-person singular pronouns (0.0415 vs. 0.0238, *p*=1.93 × 10^−8^), first-person plural pronouns (0.0150 vs. 0.0072, *p*=8.25 × 10^−8^), and second-person pronouns (0.0808 vs. 0.0748, *p*=1.88 × 10^−2^). Additionally, relative to the first quintile of the session, therapists in the final quintile tended to speak for longer durations, measured both in terms of **raw seconds per talk turn** (7.1615 seconds vs. 4.8952 seconds, *p*=7.35 × 10^−4^) as well as the ***ratio* of therapist-to-patient seconds per talk turn** (1.879 vs. 0.938, *p*=4.95 × 10^−6^). While therapists tended to speak longer in each talk turn near the end of the session, they also tended to speak faster relative to the patient, such that the **ratio of therapist words per second to patient words per second** was higher in the last quintile relative to the first (1.1715 vs. 1.040, *p*=9.62 × 10^−3^). These results were all significant after controlling the False Discovery Rate at level α=0.05 via the Benjamini-Hochberg procedure.

The aggregate trends in therapist language highlighted above were in some cases also present in patient language, but the starting point and relative alignment (i.e., parallel, convergent, divergent) varied significantly depending on the language feature under consideration. See Figure 2 and Supplementary Table 1 for additional details.

Although therapist language appears dynamic within sessions, patient language does not always follow the same trends. Figure 2 presents therapist-patient within-session language changes organized by quintile. Therapists’ use of negative and past-oriented language decreased significantly over the course of the session (Figures 2a and 2c), while their use of future-oriented language and first-person plural pronouns increased significantly (Figures 2b and 2d). In some cases, patient and therapist language features **converged** over time (e.g., Figures 2b and 2c: therapists used significantly less future-oriented language and significantly more negative language early in the session relative to patients, but these differences disappeared later in the session). In other cases, patient and therapist language **diverged** (e.g., Figure 2d: there were no significant differences between patient and therapist use of first-person plural (“We”) pronouns early in the session, but near the end of the session therapists used significantly more first-person plural pronouns than patients). In yet other cases, therapist and patient language differed significantly but seemed neither to converge nor diverge (e.g., Figure 2a: use of past-oriented language).

### Therapist speech is responsive

Here we evaluate therapist language responsiveness, specifically the degree to which changes in patient speech patterns are associated with subsequent changes in therapist speech patterns after controlling for potential confounding factors. Out of the 78 sessions we considered, two were excluded because they exhibited non-stationarity after differencing (differencing is a common technique in time series analysis for removing macro-level trends from time series whereby differences between consecutive observations are computed and treated as the primary subject of analysis; see Supplementary Methods for additional details).^49^ Another three were excluded because the patient and/or therapist had one or more language features with zero variance. Across the remaining 73 sessions analyzed, of the 18,688 possible dyad-specific associations between patient and therapist language features (16 language features each for patient and therapist, for 73 dyads) that were tested, 303 (1.6%) were significant after controlling the false discovery rate at level α=0.05. The mean (median) number of significant associations per therapist-patient dyad was 4.2 (3.0), with the minimum number of links in a session being 0, the maximum being 16, and the interquartile range (25th percentile, 75th percentile) being (2, 5). See Supplementary Figure 3 for the distribution of the number of significant links per session. Figure 3 shows directed acyclic graphs illustrating the set of associations for a subset of the therapy sessions. As illustrated in Figure 3, while the exact combinations of significant associations describing each therapist’s accommodation patterns were almost all unique, some forms of accommodation (i.e., the therapist modulating their speech patterns in response to changes in patient speech patterns) were more common than others. The top three most frequent accommodation patterns were as follows: of 78 therapists in the sample, (1) 12 therapists significantly decreased their rate of speech (as measured by words per second) in response to increases in the patient’s rate of speech, or vice versa (mean [SD] partial correlation: -0.24 [0.069]); (2) seven therapists significantly decreased their use of personal pronouns in response to increases in the patient’s rate of speech, or vice versa (mean [SD] partial correlation: -0.27 [0.064]); (3) six therapists significantly altered the frequency with which they used phrases that demonstrate understanding in response to increases/decreases in their patients’ use of third-person plural pronouns, though we note that four therapists increased their use of such phrases in response to increased patient third-person plural pronoun use (or vice versa) while two therapists’ use of such phrases moved in the opposite direction (mean [SD] partial correlation: 0.10 [0.34]). Figure 4 presents the frequency with which certain associations between patient language features and subsequent/accommodating therapist language features appeared, across all sessions (for the sake of readability, only associations represented by at least three dyads are presented - see Supplementary Figure 2 for all associations).

### Therapists are consistent between sessions

Here we evaluate therapist language consistency across sessions. The average pairwise correlation of language patterns between therapists in our primary sample, averaged across 3,003 (78 choose 2) distinct pairs of therapists, was -0.012 (95% CI: [-0.0218, -0.0024]), while the average pairwise correlation within therapists (comparing language patterns from two sessions with the same therapist but different patients) was 0.253 (95% CI: [0.1299, 0.3794]) across 20 samples. A *t* test comparing the two distributions revealed that this difference was significant at level α = 0.05 (*t* = 4.39, *p* = 1.15 × 10^−5^), suggesting that on average, within-therapist language patterns were significantly more similar than between-therapist language patterns.

### Clinical relevance: Diagnoses and symptom severity

Here we evaluate therapist language as it relates to patient diagnosis and symptom severity. Logistic regression models trained to classify diagnosis based on therapist language patterns performed significantly better than chance in terms of accuracy on a held-out evaluation set (72.04% vs. 55.26%), with an average [95% CI] model accuracy improvement over chance (i.e., always guessing the majority class) of 16.78% [5.13%, 28.21%] (*p* = 0.008). Logistic regression models trained to classify symptom severity also performed better than chance in terms of accuracy (81.97% vs. 74.45%), though the improvement of model accuracy over chance accuracy (7.52%, 95% CI: [-2.56%, 17.95%], *p* = 0.094) was not significant at level α=0.05.

## Discussion

In this work we provide researchers a transparent computational approach for representing, measuring, and analyzing therapist language in psychotherapy without time-consuming human inspection. We apply our approach to directly measure and analyze therapist language - both individually and in aggregate, and at multiple time scales (at the level of entire sessions, session quintiles, and utterances). We examine three clinically relevant but computationally neglected aspects of therapeutic discourse analysis: therapist language <u>timing</u>, <u>responsiveness</u>, and <u>consistency</u> across five clinically relevant domains: pronouns, time orientation, emotional polarity, therapist tactics, and paralinguistic style. We demonstrate the potential clinical utility of this approach by evaluating the association between therapist language and two aspects of patient treatment: diagnosis and symptom severity. We conclude that increased use of computational language analysis of therapy will allow researchers and clinicians to transition from simply knowing what was said, to understanding what is most therapeutic^50^.

### Timing

Although therapists need to decide what to say and when to say it, the temporal sequencing of therapist language has been poorly measured^13,15^. Moreover, clinical features of interest are typically analyzed in isolation, leaving potential sequencing or interactions unexplored. Our approach puts multiple clinically relevant features in context across an entire session (Figure 1), substantiating claims from discourse analysis and linguistics that words and phrases have layered and hierarchical interpretations^50,51^. We find that prospectively identified language features (i.e., pronouns, therapist tactics, etc.) display a layered and temporally nuanced pattern that may be clinically relevant, meriting further inspection in observational or controlled studies.

Therapist-patient dyads actively adjust their speech based on emergent characteristics of the conversation^51^. Yet the specific language used by a therapist may be deployed in non-obvious ways in response to their conversation partner^46^. Our findings suggest that clinically relevant language features from each speaker appear to follow both similar and different trends between language features (Figure 2). We see evidence of multiple alignments and directions of change when therapist and patient language are directly compared. Therapist and patient language are sometimes misaligned (Figure 2a), convergent (i.e., start apart and become similar) (Figures 2b and 2c), or divergent (i.e., start similar and diverge) (Figure 2d). This finding is consistent with dyadic communication research outside of therapy, which uses related concepts such as language accommodation, adjustment, style matching, and affordances^21,50–54^. Despite a lack of harmony in concept terminology, our findings align with prior work suggesting that complex linguistic interactions are likely playing out during therapy. For example, in a study of romantic couples’ texting patterns, couples’ language converged over time towards a plateau, suggesting some normative or optimal level of linguistic alignment in romantic relationships^54^. Of note in our work, some language features converged, while others diverged, suggesting an opportunity for hypothesis generation and testing of language accommodation in psychotherapy^51,55^. For example, is emotional language convergence or divergence related to patient symptom improvement? Well-powered clinical trials or naturalistic data repositories would help discern which patterns are most associated with clinical effectiveness.

### Responsiveness

Therapist responsiveness to a patient’s personal experience is a crucial difference between in-person therapy and more easily accessible mental health treatments such as bibliotherapy or internet-delivered treatment^56^. Despite the importance of patient language in therapy discourse analysis, the moment-to-moment association of therapist and patient language has been difficult to operationalize. Our findings suggest a non-obvious and complex relationship between therapists’ and patients’ language features (Figures 3 and 4). For example, it does not appear that therapists are following simple rules such as mirroring patient language and speaking style exactly, which would be relatively easy to observe and teach future clinicians to do. To understand what successful therapists are doing, more nuanced evaluation is called for. We build on prior work which often focuses on patient or therapist language in isolation, specific therapeutic approaches (e.g. motivational interviewing), or language convergence (e.g. linguistic alignment)^11,53,54,57–59^. Our findings suggest that many-to-one and one-to-many associations are playing out between therapist and patient language features.

We do not claim originality for the idea that therapist language is responsive. In early work in discourse analysis of psychotherapy, Pittenger and colleagues (1960) wrote “the details of how [language] adjustment takes place in any given instance are worth looking for… indeed, we should venture to assert that the sequential pattern of adjustment lies at the very heart of psychotherapy process^60^.” Our contribution here is a method to inspect the adjustments across features of interest in psychotherapy. Future work is needed to establish whether specific language adjustments are helpful, inert, or harmful to patients in psychotherapy.

### Consistency

If best practices are to be developed to improve therapist training and create useful markers of therapy quality, comparisons are needed across clinicians, patients, treatment settings, and time^61^. Our findings suggest some degree of linguistic stability in therapists’ use of within-session language. We refer to this as a therapist’s ‘signature’, consistent with prior work finding linguistic ‘signatures’ of emotion regulation in laboratory-based emotion regulation tasks^41^. Therapists appear to be both idiosyncratic and consistent in their use of language. Some language patterns are similar across sessions (i.e. therapist signature), while some language patterns adjust to patient or other situational factors. Therapist signatures may reflect their lived experience, preferences, or clinical training. Whether certain signatures are more clinically effective, and whether they are modifiable, is an important direction of future research. For example, some clinicians may regularly use more empathic language, a learnable skill, which may improve patient outcomes^28,62^.

### Limitations

Our study has several limitations in how features were selected; these potentially may confound variables and generalizability. **Phase 1 - feature selection**. A small group of clinicians identified clinically relevant language features based on their training and personal experience. Other reasonable people almost certainly would have made different selections. Also, our selected features do not address multilingualism or cultural variation in language use^63–66^. **Phase 2 - language evaluation**. We caution against an overly reductionist view of therapy as primarily or exclusively language based. Visual, auditory, biological, demographic, cultural, and other contextual factors may enhance, mitigate, or contradict interpretations made from language alone. We do not evaluate, nor do we claim, that therapist language always directly causes patient language or symptom improvement. It may be that patient improvement is caused by unmeasured covariates, or that therapist language is responsive to patient improvement or decompensation. Other approaches exist for feature implementation and should be evaluated, especially in the context of accuracy and appropriateness across demographic and clinical patient characteristics^43,67–71^. **Phase 3 - clinical relevance**. Clinical symptom severity measures were gathered in a college counseling setting, and thus our findings may not be generalizable to other clinicians, patients, or treatment settings. In college counseling sites, symptom severity often ranges from mild-moderate, as is true in our sample. It is unknown whether results would differ in more patients with more severe symptoms. Additionally, the sample of 98 sessions is small relative to other AI and machine learning-based studies, reflecting a well-documented limitation in psychotherapy process research^24^.

## Conclusion

If successful, computational language analysis of entire psychotherapy sessions may address long-standing criticisms of methodological rigor in psychotherapy evaluation^15,72^. If deployed ethically and fairly, this approach could assist evaluations of treatment adherence and quality^15,73–77^. To appreciate the full diversity of expression in therapy, computationally-conducted, theoretically informed evaluation may be a practical necessity^14,78^. Our goal is not to reduce opportunities for clinical spontaneity and improvisation but to develop methods to learn from skilled therapists. Our results suggest that therapist language timing, responsiveness, and consistency demonstrate patterns that merit more rigorous inspection across populations and contexts.

## Methods

### Study design

This is a retrospective cohort study of patient-therapist dyads that uses psychotherapy transcripts gathered from a completed clinical trial. The original study objectives, methods, and results have been published previously^48,79^. Written informed consent was obtained per protocol in the original trial from both patients and therapists. The study presented here was designed and conducted independently of the original clinical trial’s primary objectives. Our study had three phases: feature generation, feature measurement, and clinical relevance. In **Phase 1 (feature generation)**, our team used a modified Delphi approach to generate a list of clinically relevant language features related to therapist skill (authors ASM, BA, SA, NS)^80^. This feature list was refined based on its ability to be implemented by an expanded team of clinicians, informaticists, and computer scientists (authors ASM, SF, JF, TA, JH, AH, NS). Each feature was then implemented based on prior research and researcher judgment (authors ASM, SF). Features were selected that maximized reproducibility and transparency^81^. In **Phase 2 (feature measurement)**, features were measured and standardized for therapists and patients in 98 professionally transcribed psychotherapy transcripts. Each transcript represents a unique patient-therapist dyad. We quantitatively assessed the structure of therapist and patient language. To evaluate timing, we measured the occurrence and frequency of the clinically relevant language features noted above (grouped into pronouns, time orientation, emotional polarity, therapist tactics, and paralinguistic style) in full therapy sessions. To evaluate responsiveness, we evaluated whether changes in therapist language were associated with immediately preceding utterance-level changes in patient language. To measure consistency, we tested whether or therapists have a consistent linguistic signature across sessions with different patients. In **Phase 3 (clinical relevance)**, the relationship between therapists’ language and patients’ clinical presentation (i.e., diagnosis and symptom severity) was evaluated. The study protocol was approved by the Institutional Review Board at Stanford University.

### Dataset

Audio recordings of psychotherapy were collected per protocol during a randomized controlled trial^79^. The sessions took place between April 2013 and December 2016 at 24 college counseling sites across the United States. Non-directed counseling was offered to participants presenting with symptoms of depression or eating disorders. Transcripts were created using professional human transcriptionists; details are provided in prior work^14^. For the current study, a convenience sub-sample of unique therapist-patient dyads was selected, yielding 78 session transcripts. For therapists with more than one patient or session in our sample, a single session was randomly selected. Thus, our primary sample had 78 sessions, across 78 unique therapists and 78 unique patients. A secondary sample added an additional 20 sessions, each of which represented a second session from a therapist in the primary sample but with a different patient relative to the first. Unless otherwise explicitly stated, any analyses are with respect to the primary sample of 78 unique therapist/unique patient sessions.

Diagnosis was made by the treating clinician during the original clinical trial using the DSM-IV diagnostic criteria. Depression symptom severity was measured at the start of each session using the Patient Health Questionnaire-9 (PHQ-9), a common and validated measure of depression severity^82–84^.

### Phase 1: Feature generation

Due to a lack of validated clinical ontologies for psychotherapy, we first identified clinically relevant features using a modified Delphi approach^80^. Features reflect either clinically important constructs (e.g. emotions) or paralinguistics (e.g. rate of speech). Features were manually clustered into five domains based on prior research and clinical judgment: pronouns, time orientation, emotional polarity, specific tactics, and paralinguistics.

### Pronouns

The Linguistic Inquiry and Word Count (LIWC) program is a validated lexicon containing psychologically meaningful categories of words and word stems, including categories for various kinds of personal pronouns^32^. Our “Pronouns” features represent the number of matches between spoken words and terms in the relevant pronoun-specific LIWC category.

### Time Orientation

Time orientation of patient and therapist language is a key focus of research in mental health^38,39^. Each “Time Orientation” feature represents the number of times a word/word stem from a relevant time orientation lexicon in LIWC appears in speech^32^.

### Emotional polarity

Emotions are important in most clinical psychology theoretical orientations^34,36,37,40^. Nevertheless, there is strong disagreement on how to measure emotionality^43,85^. We chose to use the NRC Emotion Lexicon (EmoLex) to measure whether a word conveyed positive or negative sentiment because of its expansive coverage (14,182 unigrams/words) and inspectable approach, rooted in a crowdsourced layman’s understanding of each word. The “Positive emotionality” feature represents the number of words considered to have positive polarity, and similarly for the “Negative emotionality” feature.

### Therapist tactics

We used small, non-exhaustive lexicons to detect two clinically important but rarely measured therapist tactics: active listening and non-judgmental stance, adapted from prior work^21^. Active listening entails speech acts that seek to validate the patient, clarify meaning, or direct the patient towards useful experiences^86^. A non-judgmental stance is created and maintained in many ways, but one approach is to avoid absolutist language (e.g. “always”, “never”)^87^. See Supplementary Methods for additional details.

### Paralinguistic style

The meanings of words are influenced by how the words are said^88,89^. We focus on paralinguistic aspects of speech that can be measured using only transcripts. We measured the seconds taken by each therapist per talk turn, with talk turn boundaries delineated by a change in speaker in the transcript. We additionally measured therapists’ rate of speech by dividing the number of therapist-spoken words by the amount of time that the therapist spoke, as indicated by the time stamps in the transcripts. We also measured the therapist-to-patient ratio of both seconds taken per talk turn and words spoken per second. Including these ratios provides insight into whether the therapist was speaking faster or slower than the patient, as well as taking more time in each talk turn compared to the patient.

### Phase 2: Feature implementation

#### Temporal aggregation and granularity

In addition to analyzing therapist language at the level of talk turns/utterances, we aggregated features (1) at the level of session quintiles (e.g., the first 20% of the session, by time), and (2) at the entire session level. We indicate which level of aggregation was used in each subsection of the methods. There is no standard approach, and prior work has used both quintiles and deciles to segment discourse analysis^21,33^. We analyzed sessions at the level of quintiles to reduce the variance of aggregate language feature statistics within each time window while nevertheless providing sufficient temporal granularity so as to make meaningful deductions about changes in language use over time.

#### Evaluating whether therapist speech dynamically changes throughout the session

To represent therapist language, we calculated the average value of each language feature within each quintile of therapist speech. For count-based lexicon-matching features, we calculated the proportion of total words that matched a term appearing in the associated lexicon for each quintile.

To qualitatively analyze the dynamic nature of therapist language over time, we fit a natural cubic spline to the data represented by ordinally indexed session quintiles (independent variable) and quintile-aggregated language features, averaged across therapists (dependent variable)^90^. This procedure was also performed for individual therapist language features to additionally highlight heterogeneity in the way therapist language changes over time. See Figure 1 and Supplementary Figure 1.

We also quantitatively assessed patterns in therapist language features over time. For each language feature, we compared the distribution of that therapist language feature in the first and last quintile of therapy sessions, using a nonparametric Mann-Whitney U test to test for significant differences in distribution between the two quintiles^91^. Within the first and last quintiles, we also analyzed differences between patient and therapist language features using the Mann-Whitney U test. We used the Benjamini-Hochberg procedure to control the False Discovery Rate (FDR) at level α = 0.05. See Figure 2.

#### Evaluating whether therapist speech is responsive

Here we describe how our therapist language representations were used to analyze individual therapist’s accommodation patterns at the level of utterances. To better answer the question of how therapists adapt their language to patient language, we leveraged recent methodological advances in time series causal discovery for dynamical systems to identify temporal dependencies between patient and therapist language features^92^. The algorithm we employed, PCMCI, applies momentary conditional independence (MCI) tests to identify temporal links between variables, accounting for potential observed confounding. PCMCI has been shown to identify such links in observational data with good statistical power and low Type I error. For each therapist, we used PCMCI with partial correlation to identify significant links between patient language and therapist language.

Patient-to-therapist associations were recorded as significant if the associated MCI test was significant at level α = 0.05, after controlling the FDR with the Benjamini-Hochberg procedure. We additionally calculated and reported the frequency of each type of association across all sessions.

### Phase 3: Measuring clinical relevance

We next describe our approach to differentiating *between* therapy sessions. By aggregating therapists’ language features over the entire time course of the session, we obtain a 16-dimensional vector for each therapist (i.e., the therapist’s linguistic “signature”). We sought to examine: (1) whether a therapist’s “signature” is consistent across patients, over and above chance; and (2) whether these “signatures” are associated with clinically relevant patient variables, namely symptom severity and psychiatric diagnosis.

To answer (1), we calculated the cross-therapist “signature” correlations between all pairs of therapists in our primary sample, then compared that distribution to the distribution of “signature” correlations within therapists but across different patients. We used a *t-*test to test whether there were any differences in the distribution of correlations between the two groups.

To answer (2), we performed two predictive analyses via logistic regression, treating the therapist “signatures” as independent variables and the patient symptom severity classification (admitting PHQ-9 < 10 vs. PHQ-9 ≥ 10) and admitting diagnosis (depression vs. eating disorder) as the dependent variables, respectively. We randomly divided our dataset into two equally sized halves, trained a logistic regression model on one half, and evaluated the model’s accuracy on the second half. The test accuracy on the second half was compared to chance, which in this case we defined as always predicting the majority label of the dependent variable in the subsampled evaluation dataset. The difference between our model’s accuracy and chance accuracy was recorded, and this process was repeated 1000 times, using random splits of the data each time. We used the resulting distribution of accuracy differences to estimate the probability that our logistic regression model would perform no better than chance, defining a significant result as *p* < 0.05.

## Data Availability

The dataset is not publicly available due to patient privacy restrictions but may be available from the corresponding author on reasonable request.

## Code availability

The code used in this study can be found at: (link to publicly available repository be updated prior to publication)

## Acknowledgements

A.S.M. was supported by grants from the National Institutes of Health, National Center for Advancing Translational Science, Clinical and Translational Science Award (KL2TR001083 and UL1TR001085), the Stanford Department of Psychiatry Innovator Grant Program, and the Stanford Human-Centered AI Institute. S.L.F. was supported by a Big Data to Knowledge (BD2K) grant from the National Institutes of Health (T32 LM012409) and a National Defense Science and Engineering Graduate Fellowship from the Department of Defense. N.H.S acknowledges support from the Mark and Debra Lesli Endowment for the Program for AI in Healthcare at Stanford Medicine. The content is solely the responsibility of the authors and does not necessarily represent the official views of the National Institutes of Health. We thank Fei-Fei Li and G. Terence Wilson for project guidance. We thank members of 2018 Spring AI Bootcamp, Pranav Rajpurkar, Andrew Ng, Suvadip Paul, Ben Cohen-Wang, Matthew Sun for collaboration. We thank all of the participating counseling centers, directors, therapists, and student patients.

## Author contributions

A.S.M. and S.L.F contributed equally as co-first authors. A.S.M., S.L.F, A.H., B.A.A., W.S.A., N.H.S., J.A.F. conceptualized and designed the study. A.S.M., S.L.F., A.H., J.A.F., B.A.A., J.H., and N.H.S. acquired, analyzed or interpreted the data. A.S.M., S.L.F., A.H., J.A.F., B.A.A., and N.H.S. drafted the manuscript. All authors performed critical revision of the manuscript for important intellectual content. S.L.F., A.H., and J.A.F. performed statistical analysis. B.A.A. and N.H.S. provided administrative, technical, and material support. B.A.A., W.S.A., and N.H.S. supervised the study. A.S.M, S.L.F, and N.H.S. had full access to all the data. A.S.M. and S.L.F. take responsibility for the integrity of the data and the accuracy of the data analysis.

## Ethics declarations

All authors declare no competing interests.

## Supplemental information

Additional tables and figures can be found in the supplementary material.

