## Supplement for "Uncovering the linguistic characteristics of psychotherapy: a computational approach to measure therapist language timing, responsiveness, and consistency"

### Supplementary materials

**Supplementary notes** **2**
Dataset 2
Phase 1: Feature generation 2
Therapist tactics 2
Paralinguistic style 2
Phase 2: Feature implementation 3
Evaluating whether therapist speech is dynamic 3
Evaluating whether therapist speech is responsive 3
**Supplementary Figure 1.** **5**
**Supplementary Figure 2.** **6**
**Supplementary Figure 3.** **7**
**Supplementary Table 1.** **8**

### Supplementary notes

#### Dataset

For feature extraction, including the identification and counting of lexicon-specific matches within a sentence, transcribed text was converted to lowercase and punctuation other than apostrophes (e.g. “it’s” or “you’re”) were removed. For details of our transcription process see prior work (Miner et al., 2020)[14].

#### Phase 1: Feature generation

##### Therapist tactics

*Active Listening*: One approach is to express understanding through simple utterances such as ‘yes’, ‘uh-huh’, or ‘mmhmm’. These utterances are often used as continuers, statements which interrupt the patient but are meant to signal the patient should continue speaking. Hedging is a way for the therapist to state back to the patient something they heard or believe but in a way that invites patient feedback or re-interpretation (e.g. ‘it sounds like’, ‘maybe’). Hedging and checking for understanding are related to listener skill in non-therapy settings.
*Non-judgmental Stance*: By using more balanced and contextual language (e.g. ‘sometimes’, ‘often’, ‘rarely’), the therapist attempts to model a more adaptive speaking and thinking style. Absolutist language, which has not been assessed previously in therapist language, is related to anxiety and depression in non-therapy settings[87].

##### Paralinguistic style

We measured the seconds taken by each therapist per talk turn, with talk turn boundaries delineated by a change in speaker in the transcript. Because time in each session was recorded by human transcribers at the level of seconds, some of the therapists’ talk turns (8,978 or 0.19%) had an unspecified length of time less than one second. For the purpose of calculating words per second and seconds per talk turn statistics, we imputed the length of these therapist utterances to be 0.5 seconds. We additionally clipped/winsorized the number of seconds for each talk turn to be no more than 120 seconds (2 minutes) and the words per second in each talk turn to be no more than 5 words per second (5 words per second, or 300 words per minute, is approximately the rate of speech of professional auctioneers). Less than 0.1% of talk turns were affected by either form of clipping. We additionally measured therapists’ rates of speech by dividing the number of therapist-spoken words by the amount of time that the therapist spoke, as indicated by the time stamps in the transcripts. In addition to measuring paralinguistic features of the therapist independent of the patient, we also measured the therapist-to-patient ratio of both seconds taken per talk turn and words spoken per second. Including these ratios provides insight into whether the therapist was speaking faster or slower than the patient, as well as taking more time in each talk turn compared to the patient.

#### Phase 2: Feature implementation

##### Evaluating whether therapist speech is dynamic

Quintile values were given as the midpoint of the quintile bin (i.e., the independent variable value representing the first quintile was 0.1 for all therapists, 0.3 for the second quintile, and 0.9 for the last quintile). The interpolated feature values (dependent variable) were centered and scaled so that temporal trends from distinct features could be meaningfully compared side by side (see Figure 1a).

##### Evaluating whether therapist speech is responsive

While the analyses of statistical differences between therapist and patient language features within and across time can yield provocative hypotheses about the way in which therapist and patient language are related, such findings are associative and not causal. The temporal structure of our data allows for more robust inference compared to purely associational approaches (because we can reasonably assume that only past patient and therapist language influence present therapist language at any point in the psychotherapy session), but the observational nature of our data would mandate several assumptions to make the discovered associations causally valid. As we cannot necessarily validate these assumptions, we restrain from making any causal claims. These assumptions include causal sufficiency (that all causal drivers are observed in the data), faithfulness (that all observed conditional independencies are encapsulated within the learned graphical structure), and stationarity (that the distribution of the time series in consideration does not change over time). While the first two assumptions (causal sufficiency and faithfulness) are difficult or impossible to validate, especially in the absence of strong and validated theory characterizing causal mechanisms, we explicitly tested against nonstationarity for each feature and removed from our analysis those subjects whose preprocessed language features exhibited nonstationarity.

More specifically, we preprocessed our data via differencing, i.e., by representing the therapist (or patient) language features associated with a talk turn at time *t* as the raw language feature at time *t* minus the raw language feature at time *t* - 1. This approach is commonly employed in temporal causal modeling to achieve stationarity and preserves the intuition of what we would hope to measure via our causal inference approach (i.e., “do sudden increases in a particular patient language feature cause associated increases or decreases in corresponding therapist language features?”). Using the Augmented Dickey-Fuller (ADF) unit root test and Kwiatkowski-Phillips-Schmidt-Shin test for stationarity, we tested the time series distributions of each patient and therapist’s language features to assess whether they were stationary. Out of 2496 language feature distributions tested (16 language features across 78 unique sessions, with one patient and one therapist in each session), only two specific language feature distributions were identified as being nonstationary, testing at confidence level $\alpha = 0.05$. These sessions were excluded from the temporal association discovery analysis.

### Supplementary Figure 1.

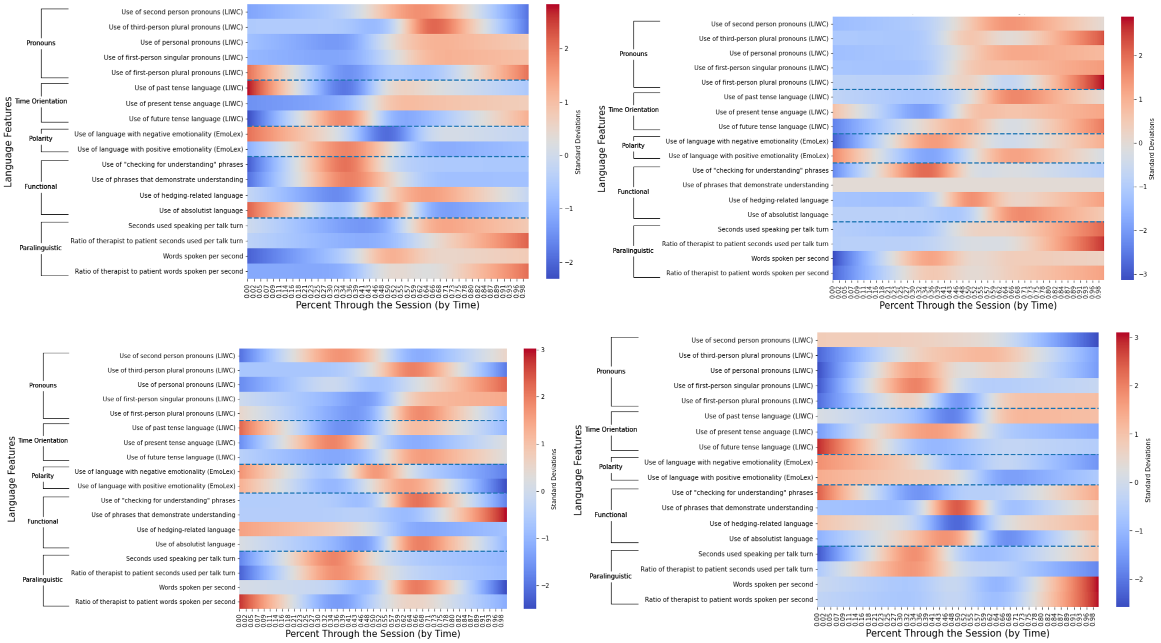


**Supplementary Figure 1:** The four figures present the same analytic approach as Figure 1 in the main manuscript, broken out into four single therapy sessions from four individual therapists.

### Supplementary Figure 2.

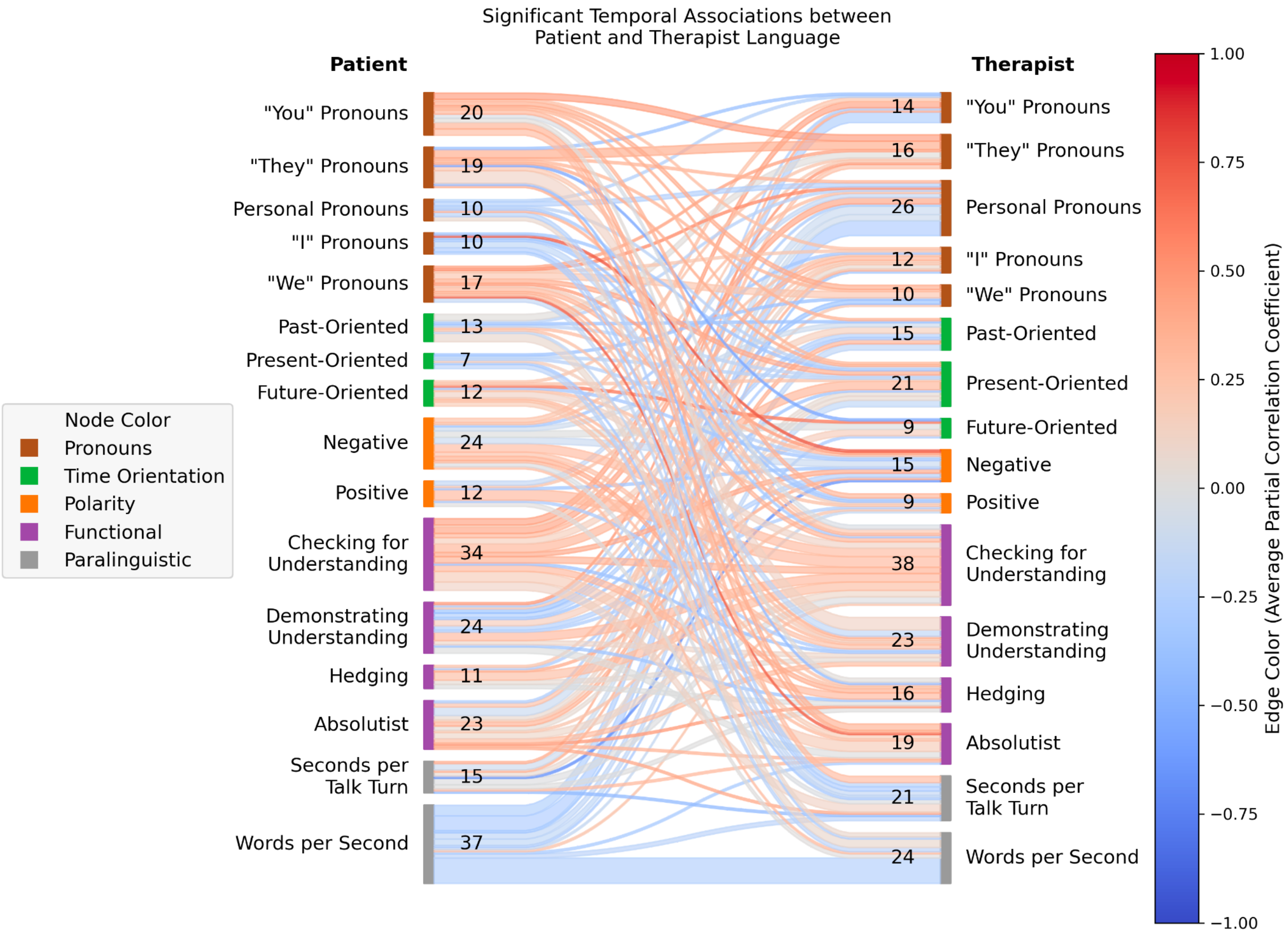


**Supplementary Figure 2:** The frequency with which certain temporal associations between patient language features and subsequent/accommodating therapist language features emerged, across all sessions. Compare to Figure 2 which illustrates a subset of edges, each of which appeared in at least four unique sessions.

Supplementary Figure 3.

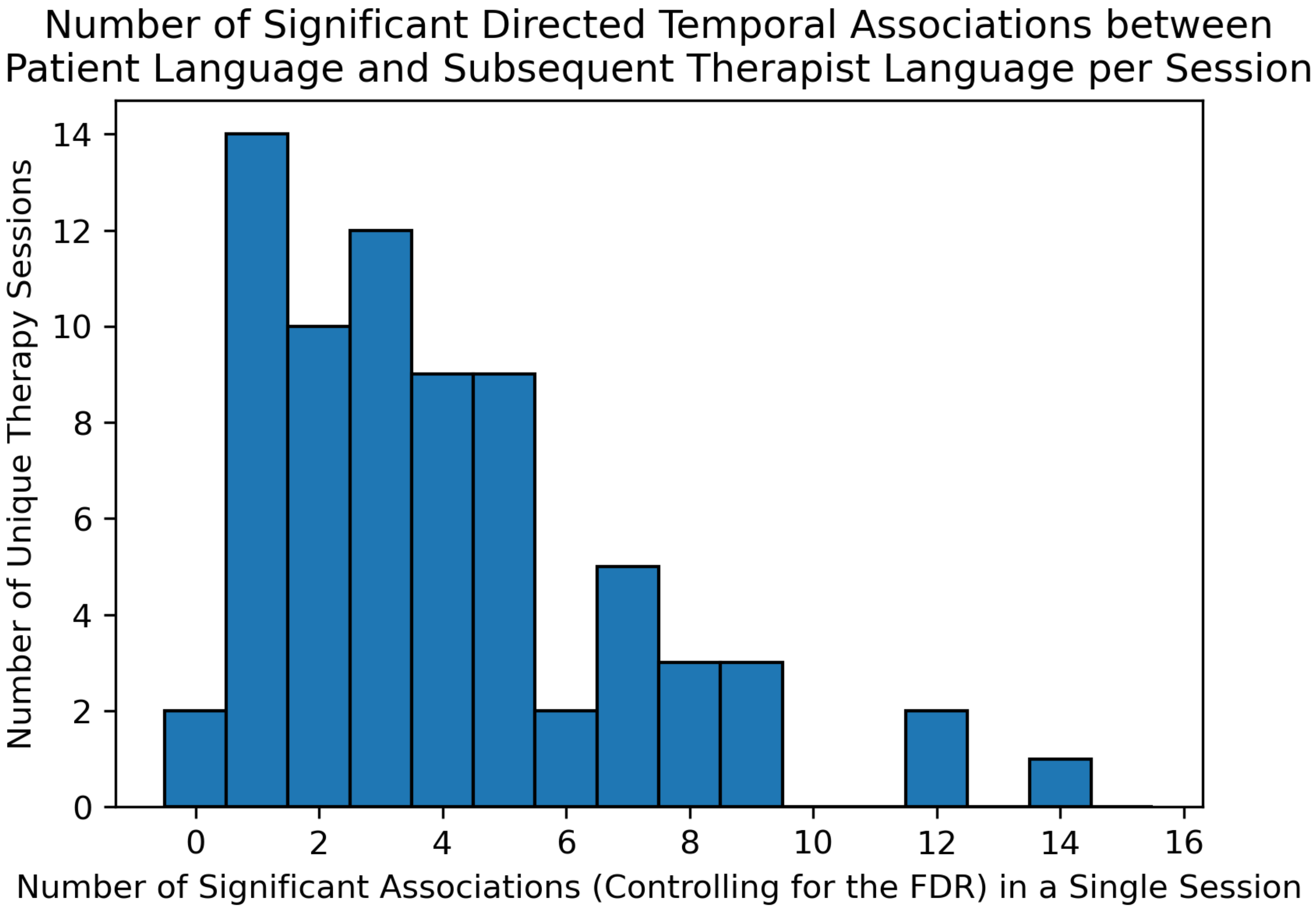


**Supplementary Figure 3:** The number of temporally dependent links between patient language and subsequent therapist language, across all sessions. For example, two sessions exhibited no significant associations and one session exhibited 14.

### Supplementary Table 1.

| | Q1 Therapist "Q1T" | Q5 Therapist "Q5T" | Q1 Patient "Q1P" | Q5 Patient "Q5P" | *P*(Q1T = Q5T) | *P*(Q1P = Q5P) | *P*(Q1T = Q1P) | *P*(Q5T = Q5P) |
|---|---|---|---|---|---|---|---|---|
| 2nd Person Pronouns | 0.074808 (0.070093, 0.079978) | 0.080765 (0.076676, 0.085386) | 0.009968 (0.008233, 0.011619) | 0.018042 (0.015310, 0.020881) | 1.88E-02 | 1.36E-05 | 4.57E-27 | 2.56E-26 |
| 3rd Person Plural Pronouns | 0.004544 (0.003235, 0.006257) | 0.005576 (0.004510, 0.006787) | 0.006948 (0.005755, 0.008305) | 0.005342 (0.003947, 0.006890) | 3.54E-02 | 1.24E-02 | 4.96E-04 | 2.84E-01 |
| All Personal Pronouns | 0.118178 (0.111600, 0.125256) | 0.149979 (0.143775, 0.156455) | 0.155274 (0.149702, 0.160996) | 0.166119 (0.159442, 0.172862) | 3.86E-10 | 2.08E-02 | 3.13E-12 | 4.50E-04 |
| 1st Person Singular Pronouns | 0.023841 (0.020646, 0.027122) | 0.041500 (0.037159, 0.046123) | 0.113585 (0.107188, 0.119677) | 0.120268 (0.112819, 0.128395) | 1.93E-08 | 2.32E-01 | 6.25E-27 | 2.78E-24 |
| 1st Person Plural Pronouns | 0.007180 (0.005458, 0.008778) | 0.015046 (0.012884, 0.017540) | 0.006715 (0.005271, 0.008253) | 0.007142 (0.005417, 0.008852) | 8.25E-08 | 9.89E-01 | 6.26E-01 | 1.93E-07 |
| Past Focus | 0.041586 (0.037730, 0.046158) | 0.023111 (0.020629, 0.025710) | 0.053799 (0.048969, 0.058963) | 0.034226 (0.030026, 0.038715) | 6.87E-11 | 1.26E-07 | 5.10E-04 | 1.25E-04 |
| Present Focus | 0.127119 (0.120388, 0.132921) | 0.169680 (0.164156, 0.175435) | 0.142528 (0.135377, 0.148580) | 0.165319 (0.157966, 0.171782) | 1.30E-15 | 2.30E-05 | 3.30E-03 | 2.12E-01 |
| Future Focus | 0.013136 (0.011337, 0.014753) | 0.020847 (0.018956, 0.022743) | 0.016827 (0.015133, 0.018558) | 0.019308 (0.016615, 0.021985) | 2.46E-07 | 4.69E-01 | 3.32E-03 | 1.09E-01 |
| Negative Emotionality | 0.022703 (0.019987, 0.025372) | 0.013624 (0.011951, 0.015131) | 0.016879 (0.015043, 0.018677) | 0.014236 (0.012142, 0.016456) | 3.97E-07 | 1.53E-02 | 8.91E-04 | 8.32E-01 |
| Positive Emotionality | 0.036382 (0.033354, 0.039576) | 0.038636 (0.035793, 0.041496) | 0.028559 (0.026553, 0.030906) | 0.029216 (0.026495, 0.031958) | 3.48E-01 | 7.16E-01 | 3.40E-06 | 8.24E-06 |
| Checking for Understanding | 0.005176 (0.003737, 0.006810) | 0.003468 (0.002374, 0.004652) | 0.000206 (0.000070, 0.000397) | 0.000753 (0.000383, 0.001212) | 3.25E-01 | 3.72E-02 | 4.79E-12 | 3.83E-07 |
| Demonstrating understanding phrases | 0.000417 (0.000179, 0.000776) | 0.000602 (0.000358, 0.000882) | 0.000342 (0.000201, 0.000493) | 0.000128 (0.000045, 0.000213) | 9.78E-02 | 2.03E-02 | 2.88E-01 | 4.56E-03 |
| Hedging | 0.020788 (0.018675, 0.023064) | 0.022689 (0.020944, 0.024366) | 0.023633 (0.021048, 0.026423) | 0.028504 (0.025967, 0.031180) | 1.10E-01 | 9.04E-03 | 1.95E-01 | 3.89E-03 |
| Absolutist | 0.007045 (0.005710, 0.008628) | 0.007681 (0.006567, 0.008824) | 0.013815 (0.011383, 0.016654) | 0.009116 (0.007968, 0.010405) | 2.19E-01 | 7.48E-03 | 2.68E-07 | 4.44E-02 |
| Seconds per talk turn | 4.895247 (3.959944, 5.938566) | 7.161465 (5.676544, 9.161874) | 8.116844 (6.393563, 10.026997) | 4.874929 (4.210783, 5.614105) | 7.35E-04 | 5.60E-04 | 5.66E-05 | 9.42E-03 |
| Seconds per talk turn ratio | 0.938381 (0.766941, 1.133695) | 1.879465 (1.490541, 2.334864) | 0.938381 (0.766941, 1.133695) | 1.879465 (1.490541, 2.334864) | 4.95E-06 | 4.95E-06 | N/A | N/A |
| Words per second | 2.364974 (2.229546, 2.505802) | 2.585216 (2.448995, 2.726731) | 2.412626 (2.258546, 2.568551) | 2.337722 (2.195653, 2.483889) | 5.63E-02 | 1.99E-01 | 3.00E-01 | 2.19E-02 |
| Words per second ratio | 1.040044 (0.963403, 1.121408) | 1.171553 (1.096911, 1.253817) | 1.040044 (0.963403, 1.121408) | 1.171553 (1.096911, 1.253817) | 9.62E-03 | 9.62E-03 | N/A | N/A |

**Supplementary Table 1:** Comparison of language features between quintiles for therapist and patient. Values on the 4 leftmost columns represent average (95% CI) feature values, with confidence intervals generated via a percentile bootstrap (1000 bootstrap samples for each feature). Values in the four rightmost columns represent *p*-values resulting from running the Mann-Whitney U test, testing against the null hypothesis that language feature distributions from one partner/quantile are equal. More specifically, for two distributions $X$ and $Y$, the Mann-Whitney U test tests against the null hypothesis that, taking a random sample from $X$ and a random sample from $Y$, the sample from $X$ is just as likely to be larger than $Y$ as it is to be smaller than $Y$. Values highlighted in green are significant after controlling the False Discovery Rate at level $\alpha = 0.05$ using the Benjamini-Hochberg procedure.
